# Development of self-phenotyping tools to empower patients and improve diagnostics

**DOI:** 10.1101/2024.06.13.24308791

**Authors:** Kent Shefchek, Sonja Ziniel, Julie A. McMurry, Catherine A Brownstein, John S. Brownstein, Erin Rooney Riggs, Matthew Might, Damian Smedley, Amy Clugston, Alan H Beggs, Heather Paterson, Peter N. Robinson, Nicole A. Vasilevsky, Ingrid A. Holm, Melissa Haendel

**Author notes:** Corresponding authors: Ingrid A. Holm, MD, MPH, Division of Genetics and Genomics Boston Children’s Hospital, 3 Blackfan Circle, CLSB 15022, Boston, MA 02115; Melissa Haendel, PhD, University of North Carolina, Chapel Hill. Joint first author (Kent Shefchek, Sonja Ziniel). Joint senior author (Ingrid A. Holm, Melissa Haendel).

## Abstract

Deep phenotyping is important for improving diagnostics and rare diseases research and is especially effective when standardized using Human Phenotype Ontology (HPO). Patients are an under-utilized source of information, so to facilitate self-phenotyping we previously “translated” HPO into plain language (“layperson HPO”). Another self-phenotyping survey, GenomeConnect, asks patient-friendly questions that map to HPO. However, self-reported data has not been assessed. Since not all HPO terms are translated to layperson HPO or in the GenomeConnect survey, we created theoretical maximum-accuracy phenotype profiles for each disease for each instrument, representing the theoretical maximum performance. Both instruments performed well in analyses of semantic similarity (area under the curve 0.991 and 0.954, respectively). To explore the real-world implications, we randomized participants with diagnosed genetic diseases to complete the GenomeConnect, Phenotypr, or both instruments. For each diagnosed disease, we compared the derived disease profile to the patient-completed profile for each instrument. Profiles resulting from participant responses to the GenomeConnect survey were more accurate than to the Phenotypr instrument. The Phenotypr instrument had a tighter distribution of scores for respondents who did both instruments and was therefore more precise. We evaluated the ability of each known Mendelian disease HPO phenotype profile to retrieve the corresponding disease. We conducted interviews and generally participants preferred the GenomeConnect multiple choice format over the autocomplete Phenotypr format. Our results demonstrate that individuals can provide rich HPO phenotype data. These results suggest that self-phenotyping source of information could be used to support diagnostics or supplement profiles created by clinicians.

## INTRODUCTION

In addition to seeing a variety of physicians, patients with an undiagnosed disease spend an enormous amount of time online and in patient communities describing their symptoms and trying to find other patients like them. Patients’ self-descriptions are often granular but are not expressed in a computer-recognizable form and are rarely utilized in clinical or informatics contexts. In addition, undiagnosed disease patients also go from specialist to specialist, often with little or no care coordination between them. Yet a complete understanding of all aspects of the features that characterize a patient’s condition (“phenotyping”) is vital to make a diagnosis and critical to inform the genetic analysis that may lead to the answer. Tools and standards for genomic data analysis have advanced dramatically over the last decade [1]; by contrast, despite the critical importance of phenotyping, collecting phenotypic data has not become more standardized or less expensive [2]. One approach to collecting comprehensive phenotyping data, taken by the NIH *Undiagnosed Diseases Program (UDP)* [3–5] and the expanded *Undiagnosed Diseases Network (UDN),* is to bring the patient to a medical center for phenotyping by different subspecialists [3–5].

The *Human Phenotype Ontology (HPO)* is a structured and logically-defined vocabulary of phenotypic features encountered in human disease. It was developed to facilitate deep phenotyping [6], whereby phenotypic findings (a phenotypic profile) are captured [7]. The resulting “HPO profile” can be used to assist with identifying the most probable candidate disease, as well as to match patients with similar phenotypes/genotypes through tools such as the *Matchmaker Exchange* [8,9]. The use of semantic similarity and probabilistic models in medical applications has increased over the past years, leading to a variety of algorithms to calculate semantic similarity between phenotype profiles and thereby support improved diagnostics [10–13].

In the typical evaluation of a patient with an undiagnosed disease, the clinical team conducts a comprehensive phenotype evaluation and assembles a list of HPO terms characterizing the patient’s phenotypes. However, even this intensive process may leave important gaps. Patients often have a uniquely comprehensive knowledge of their phenotypic features, potentially because there are phenotypes that are not asked about, are not easily observed in a clinical setting, or are not shared effectively across care providers. Thus, self-phenotyping may be a valuable addition to family history, clinical evaluation, and diagnostic testing in identifying a genetic diagnosis. In addition, self-phenotyping may be especially important for individuals who do not have access to receive comprehensive phenotyping by clinicians. Not only could self-phenotyping save time and money, but it could alleviate bottlenecks in the diagnostic odyssey and, importantly, empower patients to participate in their own care.

In light of the value of patient self-phenotyping and the need to generate HPO terms, several approaches have been taken. One is to have patients answer survey questions that describe their phenotypes; the responses can then be mapped to HPO terms. This is the approach taken by GenomeConnect, the patient registry developed by the Clinical Genome Resource (ClinGen) [14]. The GenomeConnect self-phenotyping survey was developed as a broad “review of systems” to add phenotypic context to variant observations submitted to ClinVar; on this initial survey, participants are presented with a limited number of “common” phenotype terms associated with each body system, as well as an area to describe other features with free text. Additional, more focused surveys are assigned to the participant based on their answers to the initial survey. For example, if a participant indicated that they experienced seizures, they would later be assigned the GenomeConnect seizure survey, designed to collect more in-depth information about type, onset, treatment history, etc. Within GenomeConnect, phenotypic and genomic variant information is collected on each participant with the primary goal of providing the type of information needed to adjudicate variant classifications, either by ClinGen expert panels or others. However, since the GenomeConnect survey was not designed for diagnostic use, it had not been validated to assure that the HPO terms generated by the survey accurately reflect the patient’s phenotype. We sought to do this validation as a vital step to demonstrate that patients’ production of structured phenotypic data is accurate and potentially diagnostically useful [15,16]. An alternative method for self-phenotyping is for patients to directly select HPO terms corresponding to their symptoms. However, since most HPO terms are medical terms unfamiliar to patients, this requires the HPO terms be “translated” to plain language terms that a patient would be more familiar with. This is the approach taken by the Monarch Initiative [17,18], which systematically translated plain language counterparts for each of 8,164 HPO terms, although not all HPO terms are readily translatable (Figure 1); we call this the “layperson HPO”. The utility of the layperson version of HPO in self-phenotyping was unknown as it had not been computationally assessed or evaluated in patients.

**Figure 1.**
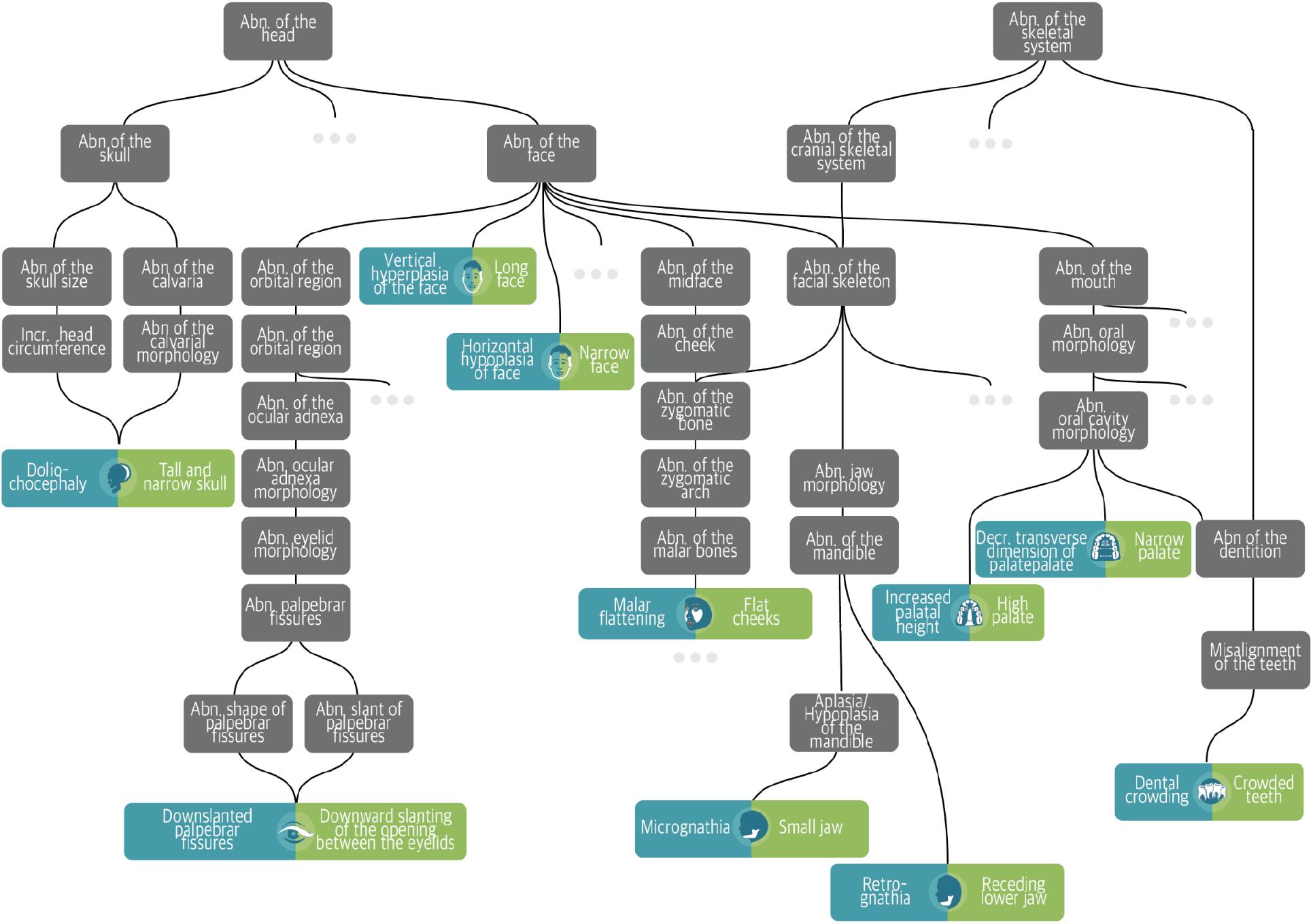
The Human Phenotype Ontology and layperson encoding. The Gold standard profile for Marfan Syndrome includes many possible facial phenotypes illustrated by the leaf nodes above. Each such phenotype is shown in its location within the overall HPO graph. For the leaf nodes, the patient-centered terms are shown in green on the right and their clinical counterparts are shown in teal to the left.

**Figure 2.**
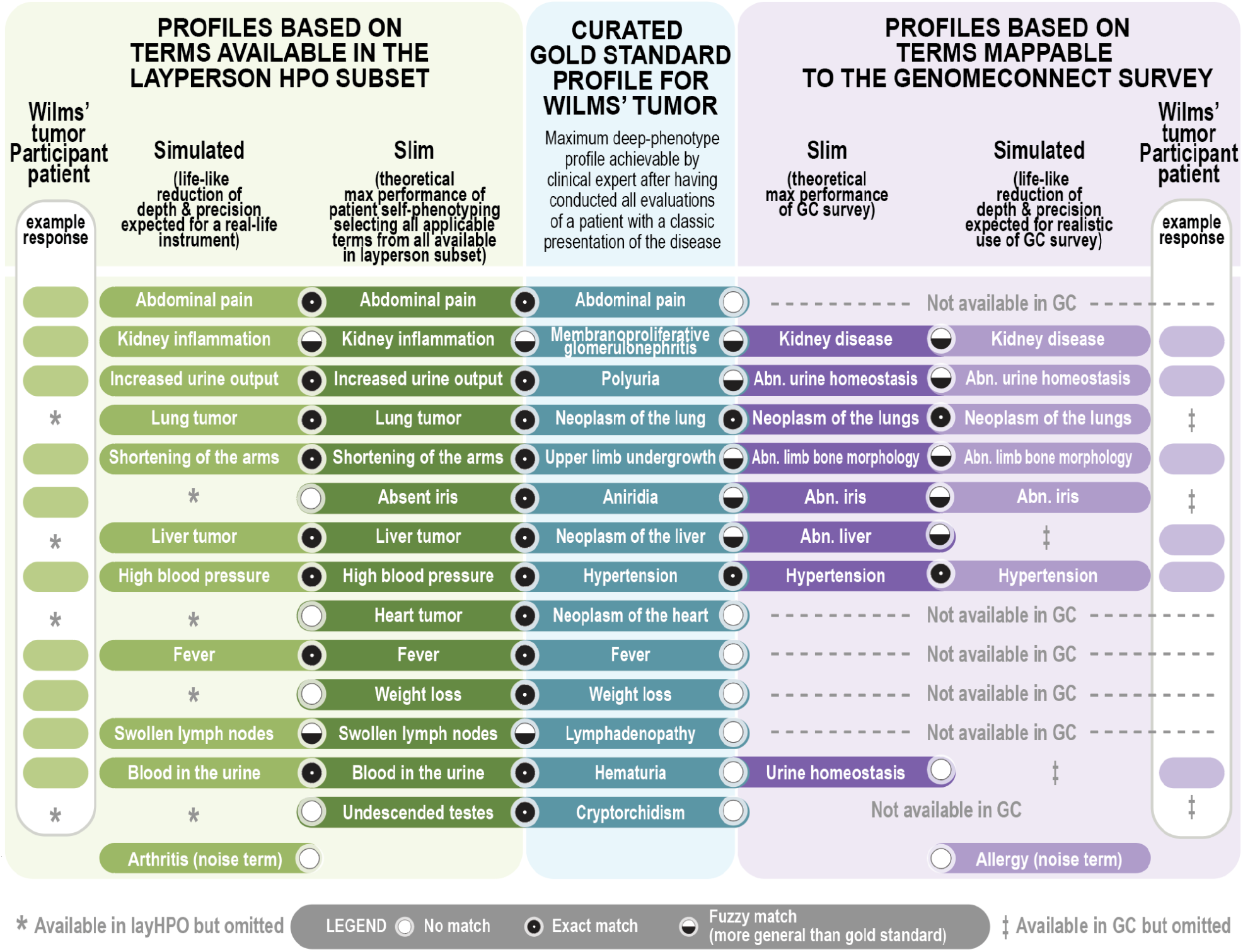
Overall phenotype profile comparison strategy (Wilms’ tumor example). Shown are example Wilms Tumor Gold standard phenotypes (teal) and their comparison to their TMax and simulated (life-like) counterparts for each of the two instruments: layperson subset of HPO (green, left) or the GenomeConnect mappings to HPO (purple, right). Example real patient responses are shown to the leftmost and rightmost columns, respectively.

The goal of this study was to assess these two self-phenotyping approaches, the *GenomeConnect survey* and the *layperson HPO* — to benchmark their theoretical maximum utility as well as to explore patients’ real-world performance. Our long-term objective is to integrate self-phenotyping into the evaluation of patients to facilitate clinical diagnostics and research.

## METHODS

### Overview of HPO profile approaches

For each of 7,344 unique known Mendelian diseases in the Monarch Initiative’s standard corpus there exists a reference phenotype profile [19] that is curated from the literature and from disease reference sources such as OMIM and Orphanet. However, not all of these clinical HPO phenotype terms are readily translatable into plain language in the layperson HPO, and only a fraction are mappable from GenomeConnect’s limited number of survey questions. We therefore sought to understand the impact that the differences in profile richness could have on the diagnostic utility of these two self-reported phenotyping approaches. For each of 7,344 known Mendelian diseases we compared the Gold standard reference HPO phenotype profile (center, teal in the **Figure 1** example, “HPO-Gold” in **Table 1**) to profiles based on terms in the layperson HPO and to profiles based on terms mappable to the GenomeConnect survey responses [20,21]. We determined, for each disease, whether the resulting HPO phenotypic profiles from the two self-phenotyping methods (the GenomeConnect survey and layperson HPO) identified that given disease after comparing against all 7,344 diseases curated with the Gold standard HPO profiles. Source profiles are available at https://github.com/monarch-initiative/hpo-survey-analysis/tree/master/data/disease_profiles.

**Table 1.** HPO profiles for each participant’s disease.

| **Table 1. HPO profiles for each participant's disease** |  |  |  |
| --- | --- | --- | --- |
|  | **Clinical-grade HPO** | **Genome Connect** | **Layperson** |
| **Gold:** Gold standard clinical HPO profile for the disease (one profile per disease) | **HPO- Gold profile** | N/A | N/A |
| **TMax:** Derived theoretical maximum profile of lay-accessible HPO terms corresponding to the Gold standard and also mappable using the instrument (one profile per disease) | N/A | **GC-TMax Profile** | **Lay-TMax Profile** |
| **Sim:** Simulated with missing terms and noise terms. HPO-Gold-Sim starts from the HPO-Gold, whereas GC-Sim and Lay-Sim start from the TMax profiles (twenty profiles per disease) | HPO-Gold-Sim | **GC-Sim Profile** | **Lay-Sim Profile** |
| **Patient:** Actual patient profile as obtained from the instrument (number of profiles dependent on patient population) | N/A | **GC-Patient Profile** | **Lay-Patient Profile** |

### Generating theoretical maximum accuracy (TMax) phenotype profiles

Because not all of the HPO is layperson accessible, and not all of the layperson HPO is available in the GenomeConnect survey, we created theoretical maximum (TMax) phenotype profiles for each of the two instruments. The TMax profiles represent the theoretical maximum patient performance for each instrument. To derive these, we started with the Gold standard reference HPO phenotype profile and omitted terms that were not available using each phenotyping method. In the case of the layperson HPO, the TMax profile is constrained to terms for which there also exists a layperson HPO synonym. In the case of GenomeConnect, the TMax profile is constrained to the HPO layperson synonyms that are mappable to questions asked in the survey. The TMax profiles therefore represent the theoretical maximum diagnostic utility achievable for a profile generated by a layperson that answers every applicable entry on each of the two instruments.

### Generating simulated “life-like” (Sim) phenotype profiles

There are many reasons patients are unlikely to encode their phenotypes to perfectly match the clinical gold standard profile for their disease, including phenotype variability, unrelated symptoms, and challenges in accurately completing the instrument. Therefore, we wanted to evaluate realistic profiles wherein we randomly added noise and made omissions. For each of the two phenotyping methods (GenomeConnect survey and the layperson HPO), we simulated 20 patients each for 7,344 rare diseases, generating 146,880 simulated profiles per method. To create each simulated patient, we started with the TMax profile based on the available HPO terms in the method by constraining the patient profile to HPO terms that had a layperson or GenomeConnect translation. We then computationally and randomly created simulated profiles from the TMax phenotype profiles by omitting terms, adding imprecision, and adding noise (defined as terms not associated with the disease). Terms that were omitted were picked randomly by the computer. Terms that were randomly selected to become less precise were replaced by their parent term. For example, if a disease was annotated to the term “Distal lower limb amyotrophy”, we would select the term “Lower limb amyotrophy” for the simulated profile. Noise was added by selecting random phenotypes in the subset not annotated to the target disease. We decided the number of terms selected based on the size of the TMax profile:10% of the terms were made less precise, 20% of the terms were omitted, and 23% of the terms in the final profile were noise.

### Permuting Gold standard phenotype profiles

For simulating profiles based on the full clinical Gold standard phenotype profiles, we increased the noise parameters since the layperson HPO and GenomeConnect derivation process naturally added omissions and imprecision. For these Gold standard simulations (HPO-Gold-Sim below), 30% of the terms were made less precise, 40% of the terms were omitted, and 23% of the terms in the final profile were noise. We ensured that all simulated profiles were unique.

### Instrument development

#### Layperson HPO

We developed an online instrument termed “*Phenotypr*” [22]. Phenotypr allows categorical selection (i.e., by anatomical system) of any layperson HPO term by the patient. There is a toggle to allow patients to include all HPO terms, and a free text box to include terms not otherwise identified.

#### GenomeConnect survey

In order to compare the GenomeConnect survey with the Phenotypr instrument, we developed a customized survey presentation platform for the GenomeConnect survey that was similar in look and feel to Phenotypr.

### Subjects

Participants were 18 years old or older, or the parent/guardian of a living child <18 years of age, diagnosed with a rare genetic disease. Participants were enrolled who were: 1) evaluated in the Boston Children’s Hospital (BCH) Genetics Clinic over the past 3 years and confirmed to have a genetic disease by the primary geneticist; 2) enrolled in the Manton Center for Orphan Disease Research at BCH and had a confirmed molecular diagnosis; 3) followed in a genetic disease-specific clinic at BCH; 4) enrolled in a patient registry for a specific genetic disease; or 5) were a member of GenomeConnect and had a confirmed genetic disease. For individuals enrolled from a disease-specific clinic, patient registry, or GenomeConnect, we confirmed their diagnosis with them prior to enrollment. Participants who completed the instrument/s were invited to enroll in the post-enrollment qualitative interviews. IRB of Boston Children’s Hospital gave ethical approval for this work (IRB-P00027106).

### Instrument distribution and data collection

Participants were offered either one or both Instruments (GenomeConnect survey and/or Phenotypr). The GenomeConnect and/or Phenotypr instruments were completed online wherever possible; a few participants for whom we did not have an email completed a paper copy of the GenomeConnect survey. Those who completed only one instrument received $15 and those who completed both instruments received $30. Those who received only one instrument were randomly assigned to the GenomeConnect or Phenotypr instrument. Participants in the Genetics Clinic were purposely assigned within a diagnosis to each method in order to assign equal numbers to each method and were matched as much as possible by gender, race, ethnicity, and age. For those who received both instruments, we alternated which instrument was assigned first. The instruments were administered via an external web interface. Each user was assigned a computer-generated unique ID. Once the data was properly entered, we captured it into the BCH instance of REDCap [23] for analysis.

### Phenotype profile semantic similarity comparisons

To assess their potential utility in a diagnostic setting, phenotype profiles were compared using semantic similarity measures. We compared the theoretical maximum (TMax) and simulated (Sim) HPO profiles for each method (GenomeConnect or Layperson HPO) against the Monarch Gold standard profiles by creating the profiles in **Table 1**.

We also compared each patient-generated profile against the corresponding theoretical maximum (TMax) profile for their diagnosed disease. Semantic similarity methods leverage the hierarchical structure of the ontology to make comparisons between graphs. A phenotype profile that is composed of ontology terms can itself be represented as a graph, each node of which represents a phenotype that corresponds to a patient or a disease (see **Figure 1** for an example phenotype profile graph representing facial features of Marfan syndrome). To calculate semantic similarity of phenotype profiles, we used PhenoDigm, an algorithm that uses both information content (Resnik similarity) and Jaccard similarity [24]. PhenoDigm takes as input two profiles, for example measuring similarity between a patient profile and a Monarch Gold standard disease profile, and outputs a score.

To run the Phenodigm algorithm, we utilized OWLTools, a java package that contains a collection of semantic similarity metrics to phenotype similarity and disease classification (https://github.com/owlcollab/owltools).

We began with full sets of TMax phenotype profiles for each instrument and the sets of simulated patients (i.e. profiles with omitted, imprecise, or noisy terms). We iterated over the phenotype profiles as input for PhenoDigm and stored the rank and similarity score of the correct disease match. The disease “rank” refers to the position of the reference disease in the prioritized list of candidate diseases suggested by the disease matching the TMax or simulated phenotype profiles against the Gold standard disease profiles. Diseases given the same score were given the same rank. Using this data, we plotted a receiving operator characteristic (ROC) curve using the Scikit-learn[25] version 0.24.2 and Matplotlib[26] version 3.4.1 python packages.

### Enrichment and depletion analysis

We hypothesized that the phenotypic profiles from the GenomeConnect survey and Phenotypr would be biased towards categories of disease with phenotypic features that are more readily visually observable (e.g., musculoskeletal disorders may be disproportionately represented in the layperson HPO terms in Phenotypr because the phenotypes are more easily observed and described in lay terms, whereas liver disease may be underrepresented because these phenotypes may not be as “visible” to the patient). In order to measure if types of diseases were over- or under-represented in each phenotype subset of HPO terms, we performed an enrichment and depletion analysis. We collected 11,113 diseases and classes of disease that had at least one phenotype term. For each disease, we generated a contingency table of the number of phenotypes in the subset of the phenotype profile associated with each disease, the number of phenotypes in the subset not associated with the disease, the number of phenotypes not in the subset associated with the disease, and the number of the phenotypes not in the subset not associated with the disease. For each disease and disease category we performed a Fisher exact test to characterize the distribution of phenotype terms across our disease phenotype profiles and adjusted the p-values with Bonferroni correction.

### Statistical Methods

We calculated effect sizes to determine when differences in methods were substantial and report the absolute value of effect sizes. For <u>quantile regressions</u> we standardized the continuous variables included so that they had a mean of 0 and a variance of 1. The resulting regression coefficients can be interpreted as effect sizes across all quantile regressions. We report absolute values that range from 0 to 1; higher numbers represent stronger effects. For <u>logistic regressions</u> we used the standardized coefficients that can be interpreted as odds-ratios and effect sizes across all logistic regressions. Odds-ratios range from 0 to infinity. The closer to 1, the weaker the effect; the closer to 0 or infinity, the stronger the effect. For <u>2×2 contingency</u> <u>tables,</u> where <u>Fisher’s exact tests</u> were used to assess the association between two variables, we also used odds-ratios. Effect sizes for equality of variance tests are not specifically developed so are not reported. The **estimated median difference** was determined by estimating the median after subtracting the GenomeConnect similarity score from the Phenotypr similarity score and was negative if the GenomeConnect score was higher (more similar) than the Phenotypr score.

Our analyses of similarity scores and ranks were all conducted separately in two groups of participants: 1) those who completed one instrument (“*One Survey*”) and 2) those who completed both instruments (“*Both Surveys*”). All analyses of the Both Surveys group accounted for the clustering/pairing of the data. For both groups we used quantile regressions to determine statistical differences between the median similarity scores, and between median ranks, between the methods. To determine if the distribution of similarity scores from one method was tighter than the other for the groups that completed both instruments, we used Pitman’s test for equality of variances; for the group that who completed only one instrument we tested for the equality of variances between the two methods using Levene’s test. To determine which method yielded more respondents where the correct clinical disease was at rank 10 or lower, for the group that completed both instruments, we generated a dichotomous variable that indicated if this was the case for a given respondent and then used the method/instrument type as an independent variable in a logistic regression; for the group that completed one instrument we used Fisher’s exact test.

### Correction for multiple testing

For these analyses, we conducted 102 statistical significance tests (not all data shown). The final p-values for all of the statistical tests were adjusted for multiple comparisons using the **Benjamini-Hochberg adjustment** as the Bonferroni adjustment is generally too conservative. Using the Benjamini-Hochberg methodology to adjust for multiple testing the new critical p-value is **<u>p<0.015</u>**. We denoted as “ns” (not significant) any values that would be statistically significant at a p-value of p<0.05 but that are no longer statistically significant due to multiple testing.

### Post-enrollment qualitative interviews

We invited all participants who completed one/both instruments to email us if they were interested in completing an interview. We asked general questions about the instrument that the participant completed, including what they thought the purpose was, the value, how they thought the instrument could be used, and if they thought the instrument would have helped in their journey to get a diagnosis. We then asked if the instrument was user-friendly, how it could have been changed to be easier to use, what was challenging or frustrating about the instrument, the time it took them to complete the instrument, if they liked the look and feel, how completing the instrument made them feel, and any additional concerns. If they completed both instruments, we asked these questions for both. The interview was audio-recorded and transcribed. We provided a $30 gift card as a token of appreciation. The team reviewed the transcripts and the interviewer’s notes and themes were identified, and discrepancies reconciled to create a catalog of the most prevalent themes. Data was analyzed using descriptive statistics and recurring patterns were identified.

## RESULTS

### Evaluation of Simulated Profiles

We measured the performance of the PhenoDigm algorithm given the three input simulations (the Monarch Gold standard simulations, layperson simulated, and GenomeConnect simulated) by plotting a ROC curve and recording the area under the curve (AUC) for each input set (**Figure 3**). This approach measures the algorithm’s ability to differentiate between diseases given three sets of input data. The full set of HPO terms (Monarch Gold standard) performed the best. The layperson subset performed second best, and the GenomeConnect subset performed the least but with an AUC of 0.954, which is considered good for many classification problems. These results were not unexpected as the layperson HPO subset is approximately a third of the full HPO and the GenomeConnect HPO subset is less than 2%; as such, diseases described with increasingly less phenotypes are expected to match Gold standard diseases with less precision.

**Figure 3.**
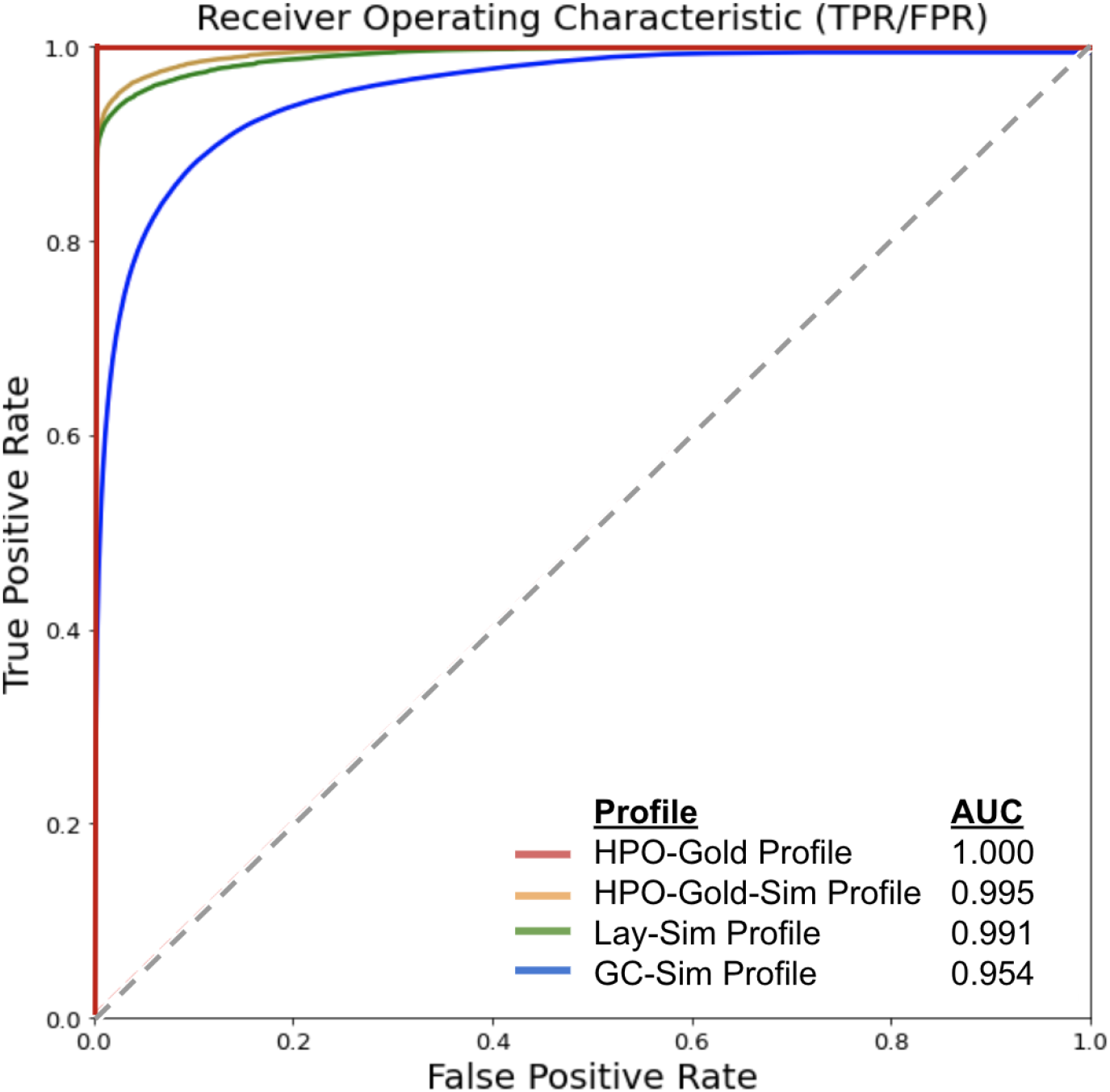
Simulated patient profiles (Lay-Sim profile, GC-Sim profile) were compared with the full HPO terminology (HPO-Gold) and its simulated counterpart (HPO-Gold-Sim). The simulations were each tested on 7,344 diseases. For each disease, 20 patients were simulated.

**Figure 4.**
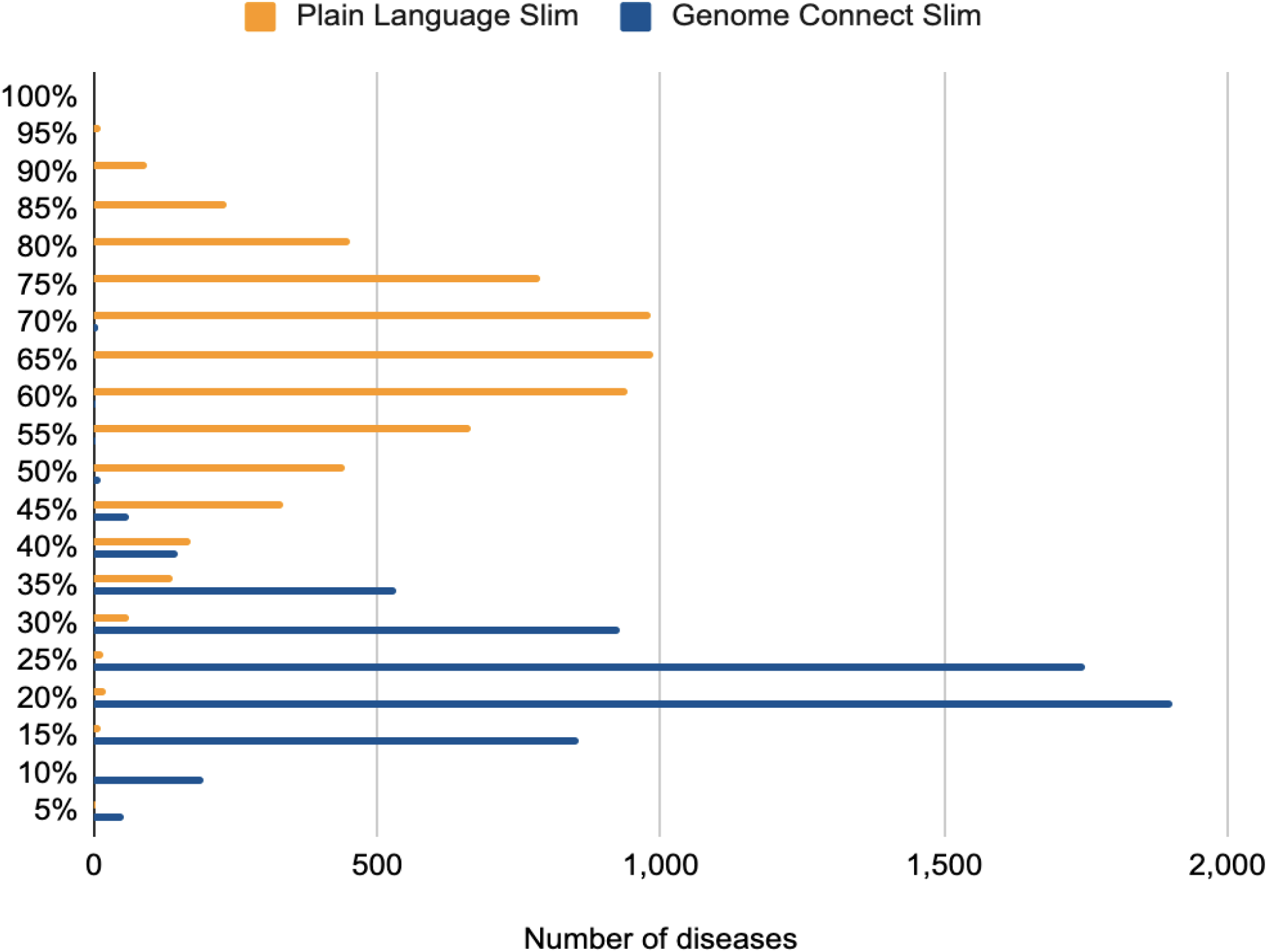
Number of diseases by percentage of lay-accessible terms. Shown are the number of diseases (x axis) and the corresponding percentage of lay-accessible HPO terms (y-axis) for each of the two instruments, where the GenomeConnect survey (blue) necessarily maps to fewer Gold standard HPO terms than does the layperson HPO available via Phenotypr (orange).

### Evaluation of 100,000 Genomes Project Cases

742 diagnosed cases from the 100,000 Genomes Project[27] were processed through Exomiser[28] (12.0.1 and 2007 databases) with default settings and either: (i) the full HPO profile collected by the recruiting clinicians or (ii) a converted profile where the terms were replaced with the closest layperson term in the hierarchy. Exomiser was able to recall 81%, 88%, and 90% of the diagnoses in the top, top 3 and top 5 ranked candidates using the full profile compared to 76%, 85%, and 87% with the converted profile, demonstrating the layHPO has sufficient coverage for use in diagnostic pipelines.

### Disease Enrichment Analysis

We used enrichment approaches to understand the classes of diseases that would be best suited to self-phenotyping approaches. We performed one-tailed Fisher’s exact tests on 11,113 diseases and classes of disease for the layperson and GenomeConnect subsets of HPO terms to test for enrichment and depletion and corrected p-values with Bonferroni correction (**Table 1**). For the layperson analysis we tested the hypothesis that, for each disease, the proportion of layperson HPO terms associated with that disease is higher than the proportion of layperson terms associated with all other diseases. We performed the same test for the GenomeConnect subset. Based on a significant p-value of p<0.01 the GenomeConnect subset was enriched for 7,480 diseases and classes of disease (and the layperson subset was enriched for 4,708 diseases and classes of disease). This suggests that there are many diseases that are significantly enriched, relative to all other diseases, with terms that a layperson could provide (via either instrument) – and therefore potentially more correctly matched by patients’ phenotyping. The source data and code are available on GitHub at https://github.com/monarch-initiative/hpo-survey-analysis/tree/master/data/enrichment.

The GenomeConnect subset of HPO terms was most enriched for vascular, neoplastic, and conjunctival diseases. The layperson subset was most enriched for bone, odontologic, and connective tissue diseases (**Table 3**). Neither the layperson nor GenomeConnect subsets had significantly depleted disease associations. Disease classes enriched with layperson terms have more phenotypes with an observable physical manifestation, such as phenotypes related to development, for example short stature, cleft lip, and short 4^th^ finger. Vascular disorders are predominantly enriched in the GenomeConnect subset, likely because GenomeConnect asks several questions about phenotypes related to vascular disorders, such as the questions “what specific types of blood/bleeding issues have you had?” and “what specific heart or blood vessel problems have you had?”.

**Table 2.** The top 10 disease terms overrepresented in the Layperson and GenomeConnect subsets. For each disease class we performed a one-tailed Fisher’s exact test on the proportion of phenotype terms in the lay person subset annotated to disease class compared to the proportion of lay phenotypes annotated to other disease classes. We performed the same analysis for the GenomeConnect subset.

| Layperson Subset Enrichment |  |  |  |
| --- | --- | --- | --- |
| Disease class | Percent of phenotype terms annotated to the <u>target</u> disease in the Lay subset | Percent of phenotype terms annotated to <u>other</u> diseases in the Lay subset | P value |
| congenital limb malformation | 59% | 28% | $1.00 \times 10^{-196}$ |
| bone development disease | 53% | 26% | $7.11 \times 10^{-195}$ |
| dysostosis | 58% | 28% | $6.27 \times 10^{-191}$ |
| bone disease | 50% | 26% | $2.04 \times 10^{-179}$ |
| rare odontologic disease | 64% | 30% | $1.91 \times 10^{-167}$ |
| skin appendage disease | 59% | 30% | $3.00 \times 10^{-161}$ |
| multiple congenital anomalies/<br>dysmorphic syndrome | 59% | 30% | $8.06 \times 10^{-161}$ |
| ectodermal dysplasia<br>syndrome | 61% | 30% | $4.26 \times 10^{-159}$ |
| eyelid disease | 57% | 30% | $1.13 \times 10^{-151}$ |
| connective tissue disease | 48% | 26% | $1.33 \times 10^{-147}$ |
| GenomeConnect Subset Enrichment |  |  |  |
| Disease Class | Percent of phenotype terms annotated to the disease in GC | Percent of phenotype terms annotated to other diseases in GC | P value |
| vascular disease | 7% | 0.30% | $1.01 \times 10^{-89}$ |
| neoplastic disease | 7% | 0.30% | $9.38 \times 10^{-89}$ |
| hypertensive disorder | 12% | 0.60% | $1.15 \times 10^{-87}$ |
| neurocristopathy | 10% | 0.50% | $6.22 \times 10^{-86}$ |
| rare adrenal disease | 11% | 0.60% | $8.02 \times 10^{-86}$ |
| conjunctival disease | 10% | 0.50% | $1.78 \times 10^{-84}$ |
| cardiomyopathy | 8% | 0.40% | $7.86 \times 10^{-84}$ |
| rare circulatory system disease | 5% | 0.20% | $5.43 \times 10^{-83}$ |
| neurovascular disease | 8% | 0.50% | $1.27E \times 10^{-82}$ |
| cardiovascular disease | 5% | 0.20% | $2.27 \times 10^{-82}$ |

**Table 3.**
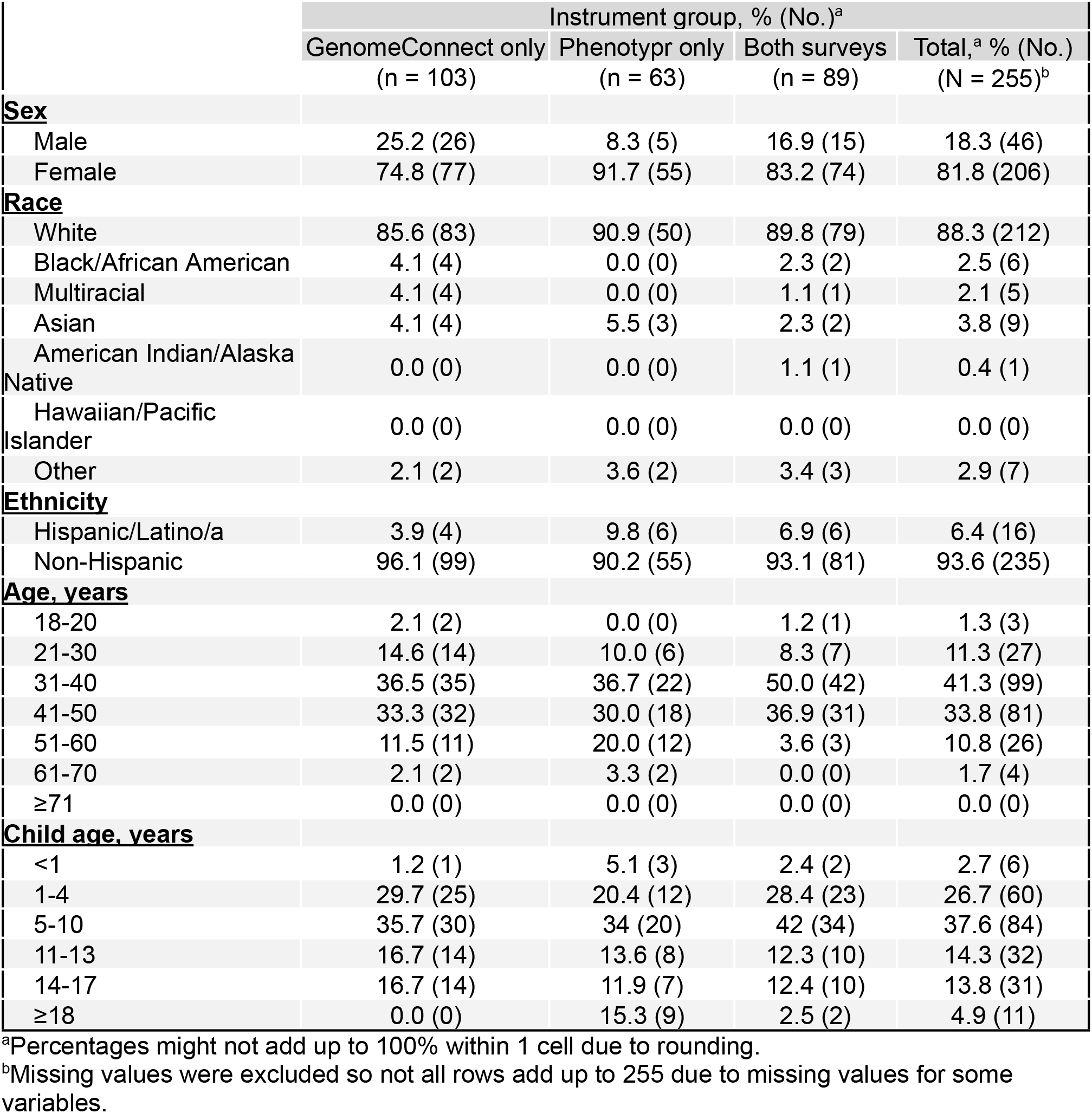
Demographic Characteristics of Final Data Set of Respondents by Instrument Group.

### Study Population

We offered enrollment to 1,061 individuals and 260 (25.0%) enrolled (**Figure 5**). The enrollment rate was 23.4% (154/659) for individuals completing one instrument, and was higher for the GenomeConnect (28.0%, 92/329) than Phenotypr (18.8%, 62/330). One participant completed the GenomeConnect survey twice — one for themselves and one for their child under the same unique identification number; this case was deleted from the dataset. We offered 402 individuals both instruments and 129 participants completed at least one (32.1%). Three of the 129 participants completed 2 instruments, one for themselves and one for their child under the same unique identification number; these 3 cases were deleted from the dataset. Of the remaining 126 participants invited to complete both instruments, 80.2% completed both (101/126), 16.7% (21/126) completed only the GenomeConnect survey, and 3.2% (4/126) completed only Phenotypr. There were 2 individuals with 2 distinct genetic diseases for which separate similarity indices could be generated. One of these individuals completed Phenotypr and one the GenomeConnect survey. In the analyses we included both similarity scores for each individual. Thus our final sample was 281 instruments and 279 respondents. Chromosomal conditions are typically out of scope for OMIM and other disease resources such as Mondo[29]; 23 respondents had chromosomal conditions and thus we removed them from the analysis. An additional respondent was dropped from the analysis because they answered all GenomeConnect questions in the negative, which was likely a user error since the phenotypic similarity algorithms use only positive phenotypes. The final dataset for analyses therefore included 257 diseases reported by 255 respondents. Of the 255 respondents, 89 answered both instruments, 63 answered Phenotypr and 103 answered GenomeConnect.

**Figure 5.**
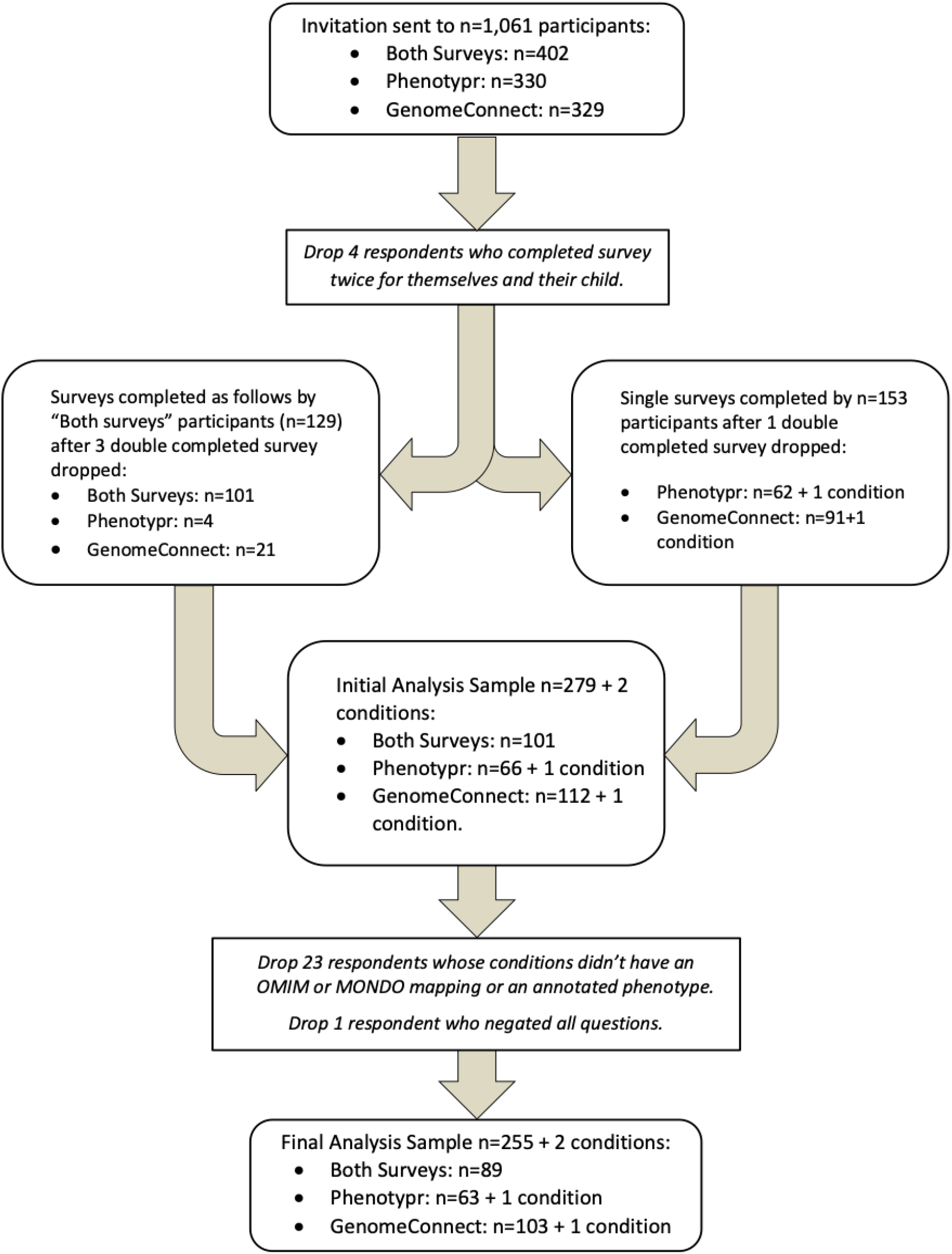
Flowchart of Recruitment, Enrollment, Survey Completion, and the Final Data Set for Analysis.

In most cases the participant was a parent completing the instrument in reference to their child who has the condition. Occasionally the respondent was an adult having the condition filling the instrument out about themselves.

Although we offered the survey to all parents regardless of race, ethnicity, or gender, respondents were predominantly white and female (**Table 4**). Most surveys were completed by the parent of a child with the condition, and the female predominance was because the mothers were generally the parent who completed the survey for their child.

**Table 4.**
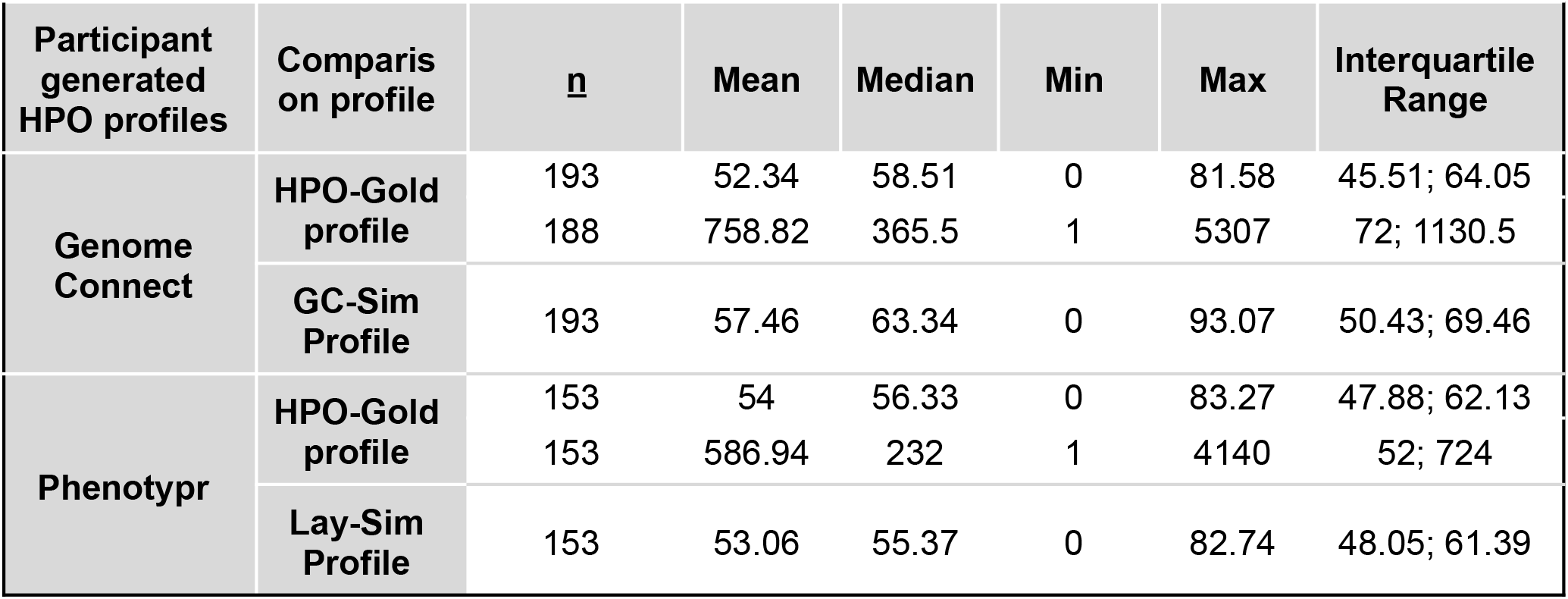
Univariate Descriptive Statistics for Similarity Scores and Ranks. Note that the n size differs because we had fewer respondents who completed the Phenotypr compared to GenomeConnect.

Profiles generated by patients with diagnosed rare diseases Univariate descriptive statistics for similarity scores and ranks are in **Table 5**.
|  |  |  |  |  |  |  |  |
| --- | --- | --- | --- | --- | --- | --- | --- |
| Participant generated HPO profiles | Comparison on profile | <u>n</u> | Mean | Median | Min | Max | Interquartile Range |
| Genome Connect | HPO-Gold profile | 193 | 52.34 | 58.51 | 0 | 81.58 | 45.51; 64.05 |
|  |  | 188 | 758.82 | 365.5 | 1 | 5307 | 72; 1130.5 |
|  | GC-Sim Profile | 193 | 57.46 | 63.34 | 0 | 93.07 | 50.43; 69.46 |
| Phenotypr | HPO-Gold profile | 153 | 54 | 56.33 | 0 | 83.27 | 47.88; 62.13 |
|  |  | 153 | 586.94 | 232 | 1 | 4140 | 52; 724 |
|  | Lay-Sim Profile | 153 | 53.06 | 55.37 | 0 | 82.74 | 48.05; 61.39 |

### Profiles generated by patients with diagnosed rare diseases

Univariate descriptive statistics for similarity scores and ranks are in **Table 5**.

**Table 5.** Analysis of GenomeConnect and Phenotypr profiles compared to the simulated profiles: Descriptive statistics of the similarity scores when compared to simulated profiles.

| Participant-generated HPO profiles | Comparison profile | One Survey |  |  | Both Surveys |  |  |
| --- | --- | --- | --- | --- | --- | --- | --- |
|  |  | n | median score | IQR | n | median score | IQR |
| GC-Patient Profile | GC-Sim Profile | 104 | 64.61 | 49.93; 69.38 | 89 | 61.69 | 50.43; 70.40 |
| Lay-Patient Profile | Lay-Sim Profile | 64 | 57.97 | 49.71; 63.31 | 89 | 54.16 | 45.73; 60.08 |

### Analysis of similarity scores

#### Comparison to simulated profiles

The descriptive statistics for the participant derived similarity scores compared to simulated profiles for both the GenomeConnect survey and the Phenotypr survey, and for the One Survey and Both Surveys groups, are in **Table 6**.

**Table 6.** Analysis of GenomeConnect and Phenotypr profiles compared to the simulated profiles: Median Similarity Scores Between Phenotypr and GenomeConnect.

|  | One Survey | Both Surveys |
| --- | --- | --- |
| Estimated Median Difference; Effect Size | -6.64; 0.34 | -7.53, 0.52 |
| 95% Confidence Interval | -1.68; -11.12 | -7.63; -7.42 |
| P value | 0.008 | < 0.001 |

For both the One Survey and Both Surveys group analyses, the median similarity score between the GenomeConnect survey profiles and the simulated profiles was significantly higher than the similarity scores between the Phenotypr profiles and the simulated profiles (One Survey: p=0.008; Both Surveys: p<0.001) (**Table 7**), demonstrating that the GenomeConnect survey HPO profiles were closer to the simulated profiles. The higher median similarity score with GenomeConnect survey suggests GenomeConnect was more <u>accurate</u> than Phenotypr.

**Table 7.** Analysis of GenomeConnect and Phenotypr profiles compared to the simulated profiles: Tightness of Distribution Between Phenotypr and GenomeConnect.

| Participant-generated HPO profiles | Comparison profile | One Survey |  |  | Both Surveys |  |  |
| --- | --- | --- | --- | --- | --- | --- | --- |
|  |  | mean | SD | p value | mean | SD | p value |
| GC-Patient Profile | GC-Sim Profile | 56.49 | 21.98 | 0.021 (ns) | 58.59 | 15.8 | 0.005 |
| Lay-Patient Profile | Lay-Sim Profile | 54.88 | 12.54 |  | 51.76 | 12.04 |  |

For the Both groups analysis only, the distribution of similarity scores between the Phenotypr profiles and the simulated HPO profiles was significantly tighter than the distribution of similarity scores between the GenomeConnect survey profiles and the simulated profiles (Both instruments: p=0.005) (**Table 8**), demonstrating that the Phenotypr profiles has less variability and thus was more precise in the Both group analysis.

**Table 8.** Analysis of Paired Rank: Descriptive statistics of disease ranks for participants who completed one survey and both surveys.

|  |  |  |  |  |  |  |  |
| --- | --- | --- | --- | --- | --- | --- | --- |
| Participant-generated HPO profiles | Comparison profile | One Survey |  |  | Both Surveys |  |  |
|  |  | n | median score | IQR | n | median score | IQR |
| GC-Patient Profile | Gold Standard | 99 | 309 | 33; 903 | 89 | 387 | 142; 1162 |
| Lay-Patient Profile | Gold Standard | 64 | 156.5 | 19.5; 613 | 89 | 261 | 89; 836 |

For respondents who completed one survey, there was no significant difference (Benjaminin-Hochberg adjusted p-value) in the distribution of similarity scores between the simulated HPO profiles and either the GenomeConnect survey HPO profiles or the Phenotypr survey HPO profiles (**Table 8**).

#### Comparison to Monarch Gold standard disease profiles

We also compared the similarity between the patient-derived profile (Phenotypr and GenomeConnect) and the Monarch Gold standard disease profiles. There were no differences in the median similarity scores between the Monarch Gold standard profiles and the Phenotypr and GenomeConnect profiles, respectively (data not shown).

### Analysis of Ranks

We compared the ranks (the position of the reference disease in the prioritized list of candidate diseases) to see which instrument identified the correct clinical disease “sooner” going down the rank list of diseases. The lower the median score, the better the rank, the closer the disease was to the top of the prioritized list of candidate diseases, and the sooner the correct clinical disease was identified. Note that due to the generality of the HPO term(s), or very few or very many terms used to describe a disease, there can sometimes be many diseases listed at the same rank.

The descriptive statistics (**Table 8**) showed that for both groups, with the GenomeConnect survey the rank had a higher median (was less accurate) compared to Phenotypr. For the One Survey group, three diseases ranked first using Phenotypr, and one using GenomeConnect (data not shown).

In the statistical comparison of the ranks, the GenomeConnect survey yielded significantly lower ranks (closer to the top of the list) for the Both Surveys group (p=0.001); but not the One group (**Table 9**).

**Table 9.** Analysis of Paired Rank: Compare Ranks Between the Methods (Phenotypr compared to GenomeConnect)

|  |  |  |
| --- | --- | --- |
|  | One Survey | Both Surveys |
| Estimated Median Difference; Effect Size | -131; 0.15 | -126; 0.13 |
| 95% Confidence Interval | -346.97; 84.97 | -131.26; -120.74 |
| P value | 0.233 | <0.001 |

There was no statistical difference between Phenotypr and GenomeConnect in the percent of participant responses that led to the correct disease ranking number 10 or better in the list of the diseases generated by participants’ responses to the instruments (correct clinical disease identified “sooner”), although there was a tendency for Phenotypr to perform better than GenomeConnect (**Table 10**).

**Table 10.** Analysis of Paired Rank: Compare Proportion of Respondents with a Rank of 10 or Lower.

| Participant-generated HPO profiles | Comparison profile | One Survey |  |  | Both Surveys |  |  |
| --- | --- | --- | --- | --- | --- | --- | --- |
|  |  | n | % rank (n) | p value; effect size | n | % rank (n) | p value; effect size |
| GenomeConnect -Patient Profile | Gold Standard | 104 | 9.62 (10) | 0.04 (ns);<br>2.69 | 89 | 2.3 (2) | 0.021 (ns);<br>4.3 |
| Lay-Patient Profile | Gold Standard | 64 | 21.88 (14) |  | 89 | 9.0 (9) |  |

### Post-enrollment qualitative interviews

#### Study Population

We conducted 17 interviews: 5 with individuals who completed the GenomeConnect survey, 5 with individuals who completed Phenotypr, and 7 with individuals who completed both instruments.

#### Themes identified

Overall, participants preferred the GenomeConnect multiple-choice format over the autocomplete Phenotypr format.

##### General themes

● The instruments will be helpful to patients/clinicians to input signs and symptoms and see suggested diagnoses.
● It was satisfying to see all signs and symptoms listed in one place.
● The length of both instruments was acceptable.

##### Phenotypr themes

- The typing aspect of Phenotypr was difficult and a list would be preferred.
- It wasn’t always clear which organ system to put a symptom under for Phenotypr.
- Phenotypr language was at times very clinical and difficult to understand

##### GenomeConnect themes

- GenomeConnect was a bit too broad (terms too general) and not granular enough.
- The layout and structure of GenomeConnect was more streamlined and manageable.

## DISCUSSION

The hypothesis of our study was that “self-phenotyping” is an accurate and comprehensive source of data on, and by, patients. Self-phenotyping is not intended to lead to a diagnosis on its own, but rather could be used as a tool to empower patients to participate in the diagnostic process, which includes the clinical evaluation, family history, laboratory work, and so forth. Although some work has gone into assessing the potential role of self-reported health data in complementing electronic health record (EHR) data [30], little has been done to assess the role and value of self-phenotyping in informing clinical care or research. In addition, self-phenotyping instruments, such as the GenomeConnect survey or use of the layperson version of HPO, have not been tested to determine if they result in accurate profiles. Further, the layperson version of HPO had not been tested as a self-phenotyping method in patients. The primary goal of this study was to determine if patients could effectively utilize the layperson version of the HPO to self-phenotype at a level that could be clinically useful, which we defined as leading to a survey-derived HPO profile that was similar to the HPO profile of the disease. We aimed to compare use of the full lay HPO in a new patient-centered application, Phenotypr, with a whole-body survey provided by GenomeConnect.

In order to address the question “can patients generate useful HPO-based data for use in disease diagnosis?”, we first created a TMax lay-subset profile (Lay-Sim Profile) corresponding to each disease in the Gold standard (HPO-Gold profile) corpus of HPO annotations. For each disease we then then compared the HPO-Gold profiles with theirTMax counterparts using semantic similarity approaches to determine “how close” the lay versions of the disease profiles would be to the correct clinical disease when compared against all diseases. This result is essentially the upper limit of what a patient might be able to achieve if they documented their phenotype profile perfectly using the full availability of the layperson HPO. The results of the similarity analysis for these TMax lay profiles showed that indeed, patients could theoretically generate diagnostically useful profiles.

We then compared simulated instrument responses (GC-Sim Profile, Lay-Sim Profile) to their HPO-Gold profile counterparts to determine how close the simulated profiles were to the Gold standard profiles. The results of the simulation showed that the layperson HPO has the capability of generating phenotypic profiles closer to the Gold standard than GenomeConnect. This is expected, since the layperson subset contains 4,757 terms in comparison to the 215 terms mapped by GenomeConnect. We concluded that Lay-Sim Profile profiles are effective at identifying the correct clinical disease, with 61% of simulated layperson HPO and 25% of simulated GenomeConnect surveys ranking the clinical disease in the top 10. We note that there are often ties based upon there being either very general HPO terms used, or very few or very many terms used to describe any given disease. This can lead to accurate, but not very precise comparisons and ranking.

We then tested the GenomeConnect survey and Phenotypr (the layperson HPO instrument that we developed) in participants with diagnosed genetic diseases to determine which performed better and which instrument was preferred by participants. The comparison with the simulated profiles allowed us to see how participants did compared to the best they could theoretically do if the instrument was perfectly completed. The comparison to the Monarch Gold standard gave us a comparison that was more clinically relevant as the clinician uses the equivalent comparison to reach a candidate diagnosis. Although the comparison to the Monarch Gold standard gave us a comparison that was more clinically relevant, since the clinician uses the equivalent comparison to reach a candidate diagnosis, the primary goal of this study was to compare the instruments themselves and assess how well participants could fill out the instruments relative to their theoretical maximum.

We found that the GenomeConnect survey had a higher median similarity score compared to the simulated HPO profiles than Phenotypr so is more **accurate**, but Phenotypr had a tighter distribution of scores so is more **precise**. However, we did not find any differences between the two instruments when compared to the Monarch Gold standard.

A key comparison is the accuracy of the instruments at identifying the correct condition. We used the rank scores to address this issue. We found a tendency for Phenotypr to be better than GenomeConnect at including the correct clinical disease in the list of the top 10 diseases generated by participants’ responses to the instruments. For many of the analyses we compared 2 groups of respondents: those who did both instruments and those who only did one. This was necessary in order to make a comparison but may have decreased our power to see differences as we divided our cohort into smaller groups. Based on the finding that Phenotypr seems to be better at identifying the disease, we conclude that Phenotypr might be more useful for clinicians to garner useful phenotype data for diagnostic use from patients. However, despite this difference, most of the time the patient’s disease was not in the top 10 diseases generated by the patient’s responses to the instruments. This suggests that more work is needed to refine the self-phenotyping strategy as well as to validate how to best utilize patient-phenotyping in the context of clinical phenotyping.

The largest conceptual difference between GenomeConnect and Phenotypr approaches is that GenomeConnect has a multiple-choice format, which our interview results showed that participants in general preferred. However, Phenotypr was more granular and able to generate a much larger number of terms for a given disease HPO profile. The responses to the qualitative interviews suggest that this granularity was overwhelming for some participants. We hypothesize that the multiple-choice format of GenomeConnect is easier for users. Phenotypr requires users to recall phenotypes, placing burden on the user to both remember and start typing a term with lexical similarity to one of the lay or clinical HPO terms. Many users provided text in the additional signs and symptoms free text field, suggesting it is likely that users were unable to find all of their phenotypes (signs and symptoms). We also hypothesize that it is easier for users to select more general terms in the HPO. For example, it is easier to say if a participant or family member experiences “behavioral abnormalities” versus if they experience “obsessive compulsive behavior”. In addition, participants found the Phenotypr language to be clinical and difficult at times. This was likely because both layperson HPO terms and medical HPO terms were included in the terms that respondents had to choose from.

Although our study advances methods for self-phenotyping, the results do not suggest a clear-cut preferred method for self-phenotyping for patients and caregivers. We conclude that a survey model that combines elements of both instruments might be the most ideal, for example, a multiple-choice survey format (GenomeConnect) that had more granular choices (Phenotypr).

In this study we validated self-phenotyping methods computationally and in a patient cohort. We have been able to demonstrate that both instruments can enable patients to provide critical phenotype information to the clinician. Ongoing work would leverage the cognitively preferred method of multiple-choice type of selection within a survey, with the more precise free-association model used when autocompleting on the lay HPO terms directly.

There are many strengths to our study. We have created a novel tool, Phenotypr, for patients to perform self-phenotyping and implemented the tools in the largest study of such tools that we are aware of. We have shown that both tools are useful in getting phenotype data directly from patients for use clinically, with the complementary strengths and weaknesses of both instruments suggesting hybrid models and/or improvements to such instruments in the future.

Our study has some limitations. The number of different diagnoses in our patient cohort was large. This meant that the robustness with respect to disease diversity was high and therefore more representative of the real-world disease heterogeneity. However, it also meant that the ability to make within-disease comparisons was challenging due to the very small cohort size for most diseases. Further, participants with diagnoses are more likely to have researched their condition and have better understanding of the clinical terms. These instruments need to be evaluated in an undiagnosed cohort to understand how they perform in a clinical setting to inform diagnosing diseases. These cohort limitations are simply the reality of working on rare diseases in general and that the diverse cohort we evaluated in the end provisioned us a reasonable spectrum of endpoints. We favored breadth over depth in any one disease for its better reflection of how the tools would be used clinically.

It should be noted that GenomeConnect was not developed for, and never meant to be, a diagnostic tool. The GenomeConnect survey is the first in a series of surveys that participants are presented with the goal of providing additional phenotypic context to ClinVar variant entries to aid in the variant classification process. GenomeConnect participants also consent to be recontacted; additional, more specific information can be obtained directly from the participant as needed and is not necessarily limited to survey answers. Thus, the survey used is intentionally broad, as it is meant to be a review of systems; more detailed surveys are assigned based on the participant’s answers. This is why there are so few terms mappable from GenomeConnect.

Finally, although we offered the instruments regardless of race, ethnicity, or gender, a limitation of this study is that the population from which we recruited was not very diverse (primarily white and female). It is therefore not surprising that mostly women (mothers) and white individuals answered.

The tools developed in this project are not yet ready for clinical diagnostic use. Future work should focus on developing an instrument that strikes the right balance between patient-friendliness and granularity. Such an instrument should be tested in a more diverse, undiagnosed disease population to determine if it leads to improved diagnostic rate or efficiency, and greater patient satisfaction in active participation of the diagnostic odyssey. Both of these outcomes are the ultimate goal of self-phenotyping. Validation and a greater understanding of how patient-phenotyping and clinician phenotyping can be best utilized together are needed. In order to address the lack of racial and ethnic diversity, reaching out specifically to underrepresented minorities would be a good strategy in future research. It would also be useful to define the literacy or education level needed to use these instruments as well. Finally, studies to determine whether patients find self-phenotyping useful or informative could be conducted, for example by surveying patients after completing the survey or conducting interviews.

In conclusion, computationally, better phenotype profiles were generated with Phenotypr. When participants completed the instruments, they preferred the GenomeConnect survey format and the GenomeConnect survey was more accurate. However, the Phenotypr instrument was more precise. This suggests that a hybrid approach that provides familiar tooling but access to richer HPO terms may be warranted. Such tools could be used to improve and accelerate diagnostic pipelines and promote collaboration and patient engagement with clinical caregivers and diagnosticians.

## Data Availability

All data produced in the present work are contained in the manuscript or are available online at https://github.com/monarch-initiative/hpo-survey-analysis/tree/master/data/disease_profiles

## ACKNOWLEDGEMENTS

The authors are grateful to Justin Ramsdill for his outstanding project management, which played a crucial role in the successful development of the Phenotypr software. This research was supported by PCORI grant HSRP20181624: https://www.pcori.org/research-results/2017/testing-two-patient-surveys-diagnosing-rare-genetic-conditions and was made possible through access to data in the National Genomic Research Library, which is managed by Genomics England Limited (a wholly owned company of the Department of Health and Social Care). The National Genomic Research Library holds data provided by patients and collected by the NHS as part of their care and data collected as part of their participation in research. The National Genomic Research Library is funded by the National Institute for Health Research and NHS England. The Wellcome Trust, Cancer Research UK and the Medical Research Council have also funded research infrastructure.

